# Review of clinical characteristics and laboratory findings of COVID-19 in children-Systematic review and Meta-analysis

**DOI:** 10.1101/2020.09.23.20200410

**Authors:** Harmeet Kaur Kharoud, Rizwana Asim, Lianne Siegel, Lovepreet Chahal, Gagan Deep Singh

**Author notes:** **Corresponding Author:** Harmeet Kaur Kharoud, MBBS, Division of Epidemiology, School of Public Health, University of Minnesota, MN, 1108 8^th^ Street SE, Apt 201, Minneapolis, MN 55414, E mail.

## Abstract

**OBJECTIVE:** To conduct a systematic review and meta-analysis to assess the prevalence of various clinical symptoms and laboratory findings of COVID-19 in children.

**METHODS:** PubMed, MEDLINE, and SCOPUS databases were searched to include studies conducted between January 1, 2020, and July 15, 2020 which reported data about clinical characteristics and laboratory findings in laboratory-confirmed diagnosis of COVID-19 in pediatric patients. Random effects meta-analysis using generalized linear mixed models was used to estimate the pooled prevalence.

**RESULTS:** The most prevalent symptom of COVID-19 in children was 46.17% (95%CI 39.18-53.33%), followed by cough (40.15%, 95%CI 34.56-46.02%). Less common symptoms were found to be dyspnea, vomiting, nasal congestion/rhinorrhea, diarrhea, sore throat/pharyngeal congestion, headache, and fatigue. The prevalence of asymptomatic children was 17.19% (95%CI 11.02-25.82%).

The most prevalent laboratory findings in COVID-19 children were elevated Creatinine Kinase (26.86%, 95%CI 16.15-41.19%) and neutropenia (25.76%, 95%CI 13.96-42.58%). These were followed by elevated LDH, thrombocytosis, lymphocytosis, neutrophilia, elevated D Dimer, Elevated CRP, elevated ESR, leukocytosis, elevated AST and leukopenia. There was a low prevalence of elevated ALT and lymphopenia in children with COVID-19.

**CONCLUSIONS AND RELEVANCE:** This study provides estimates of the pooled prevalence of various symptoms and laboratory findings of COVID-19 in the pediatric population.

## INTRODUCTION

The coronavirus disease of 2019 (COVID-19) caused by (SARS-CoV-2) was first case noted in Wuhan, China in December 2019. It has been spreading globally since then and was declared a pandemic by WHO on 11th March 2020. COVID-19 presents with a varied range of symptoms, ranging from asymptomatic to severe symptoms like acute respiratory distress syndrome, neurological and vascular involvement. It presents more severely in adults as compared with children.(1)

Data reported from various studies showed that children who test positive for COVID-19 experience a mild form of disease.(2-6) Children and younger adults without underlying lung or immunosuppressive diseases, have a much lower risk of having severe forms of COVID-19 than other age groups.(7) We aim to study the prevalence of clinical characteristics and laboratory findings in children with COVID-19 infection.

## METHODS

### SEARCH STRATEGY AND DATABASE

The Preferred Reporting Items for Systematic reviews and Meta-analyses (PRISMA) guidelines were followed while conducting this systematic review and meta-analysis. (8) The protocol was pre-registered the International Prospective Register of Systematic Reviews (PROSPERO) database (CRD42020184308).

We conducted an extensive literature search in MEDLINE via PubMed and Ovid Scopus to collect all the relevant articles about clinical characteristics and laboratory findings in COVID-19 positive children from January 1, 2020 to July 15, 2020. We used these search terms (“COVID-19” or “SARS-CoV-2” or “2019 nCoV” or “novel coronavirus”) AND (“Pediatric” or “Children” or “adolescent” or “teen” or “young’) AND (“clinical characteristic” or “Symptoms” or “Outcomes” or “laboratory findings” or “Lab’). We did not impose any language restriction.

### INCLUSION AND EXCLUSION CRITERIA

We included the following observational studies: cohort, case-control, cross-sectional, and case series that provided relevant data about clinical presentation in children and their laboratory findings. The imposed age restriction of participants in these studies was of 18 years or younger. Patients who had laboratory confirmed diagnosis of COVID-19 were included. We excluded studies with a sample size of less than 5 and those which did not provide numerical information about symptoms and laboratory findings of COVID-19 in children. The complete results from all databases used for the review were imported into a unique EndNote library upon search completion and 377 duplicated articles were removed.

### DATA EXTRACTION AND QUALITY ASSESSMENT

After removal of duplicates, two independent reviewers (HA and GS) screened the titles and abstracts of the retrieved research articles to only include studies fulfilling the inclusion criteria. The eligibility of these articles was further assessed by evaluating the full text of search results according to eligibility criteria. The same reviewers (HK and GS) then independently extracted the data into excel sheets. The studies were then evaluated and cross-checked by a third reviewer (RA). Any discrepancies were discussed until consensus was reached. Data extracted from the studies included: author, date of publication, study design, sample size, study country, and participant characteristics, including age (0-18yrs), sex, clinical symptoms, and laboratory results.

### ASSESMENT OF METHODOLOGICAL QUALITY

The methodological quality of included observational study was assessed independently by two reviewers (HA and RA) according to the Strengthening the Reporting of Observational Studies Epidemiology (STROBE) reporting guidelines.(9) The concrete content included the definition of the study design, objectives, eligibility criteria, sample size, and assessment of outcomes. The total score for assessment was 10. Studies with scores less than 4 were considered as poor-quality studies, and between 4 and 8 were considered as intermediate-quality studies, and scores more than 8 were considered as high-quality studies.

### STATISTICAL ANALYSIS

To estimate the pooled prevalence of clinical symptoms and relevant laboratory findings in COVID-19 positive children we performed random effects meta-analysis using R version 3.6.3 and the R package “meta”.(10) The pooled prevalence was estimated using generalized linear mixed models with a logit link function and can be interpreted as the median prevalence across studies. (11) Between-study heterogeneity was assessed by using Cochran Q test and I^2^ index.(12, 13) I^2^ values less than 25% were considered to represent low heterogeneity, 26% to 50% to represent moderate heterogeneity and more than 50% to represent greater heterogeneity.

Publication bias was assessed qualitatively by visual inspection of funnel plots and quantitatively by Egger’s regression asymmetry test.(10)

Univariate meta regression was used to explore whether sample size and sex of the children explained the heterogeneity between the studies.(10) P-values < 0.05 were considered statistically significant.

## RESULTS

After initial screening of the database, 1027 articles were identified (Figure 1). Of the 1027 articles screened, 650 articles were selected after excluding duplicates. After reviewing the title and abstract, 103 articles were selected for full-text assessment for eligibility according to the inclusion criteria. Finally, 37 studies with a total of 668 children met the inclusion criteria. All of the included studies were observational studies such as case series, cross-sectional and cohort studies conducted between January 1, 2020 till July 15, 2020. Most of the studies were conducted in China (89.1%) and enrolled 5 to 2135 participants with the average age ranging from 1 years to 11 years (Table 1). Although not all included studies reported laboratory findings (Table 2), most of the studies did report data on the clinical symptomatology. The number of studies that reported various clinical symptoms is as follow: fever (36/37), cough (36/37), asymptomatic status (35/37), diarrhea (19/37), nasal congestion/rhinorrhea (21/37), vomiting (14/37), headache (10/37), dyspnea (12/37) and fatigue (11/37) in COVID-19 children (Table 2). The quality score of the included studies ranged from 7 to 10 (eTable2 from Supplement).

**Table 1.**
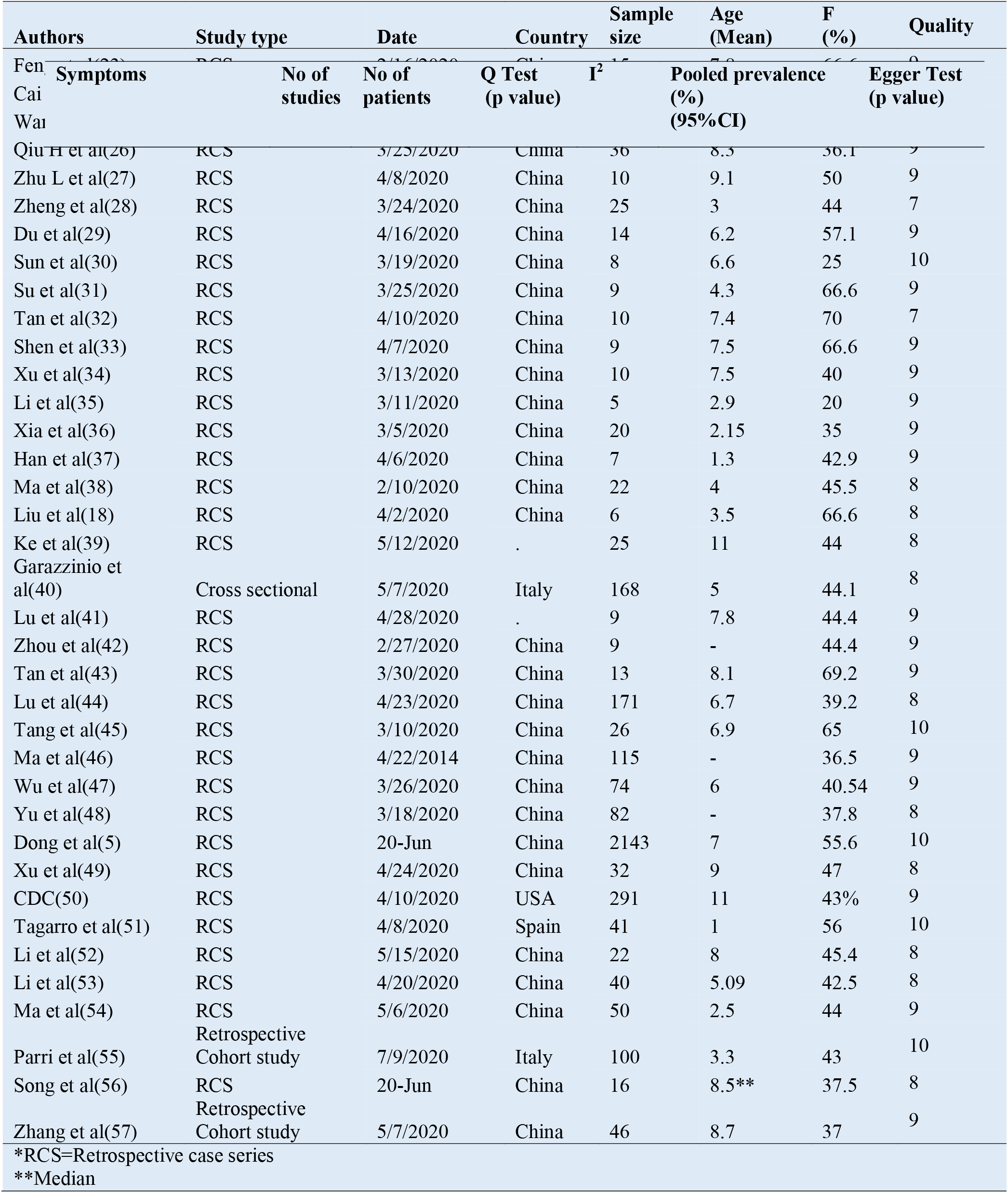
Characteristics of included studies

**Table 2.**
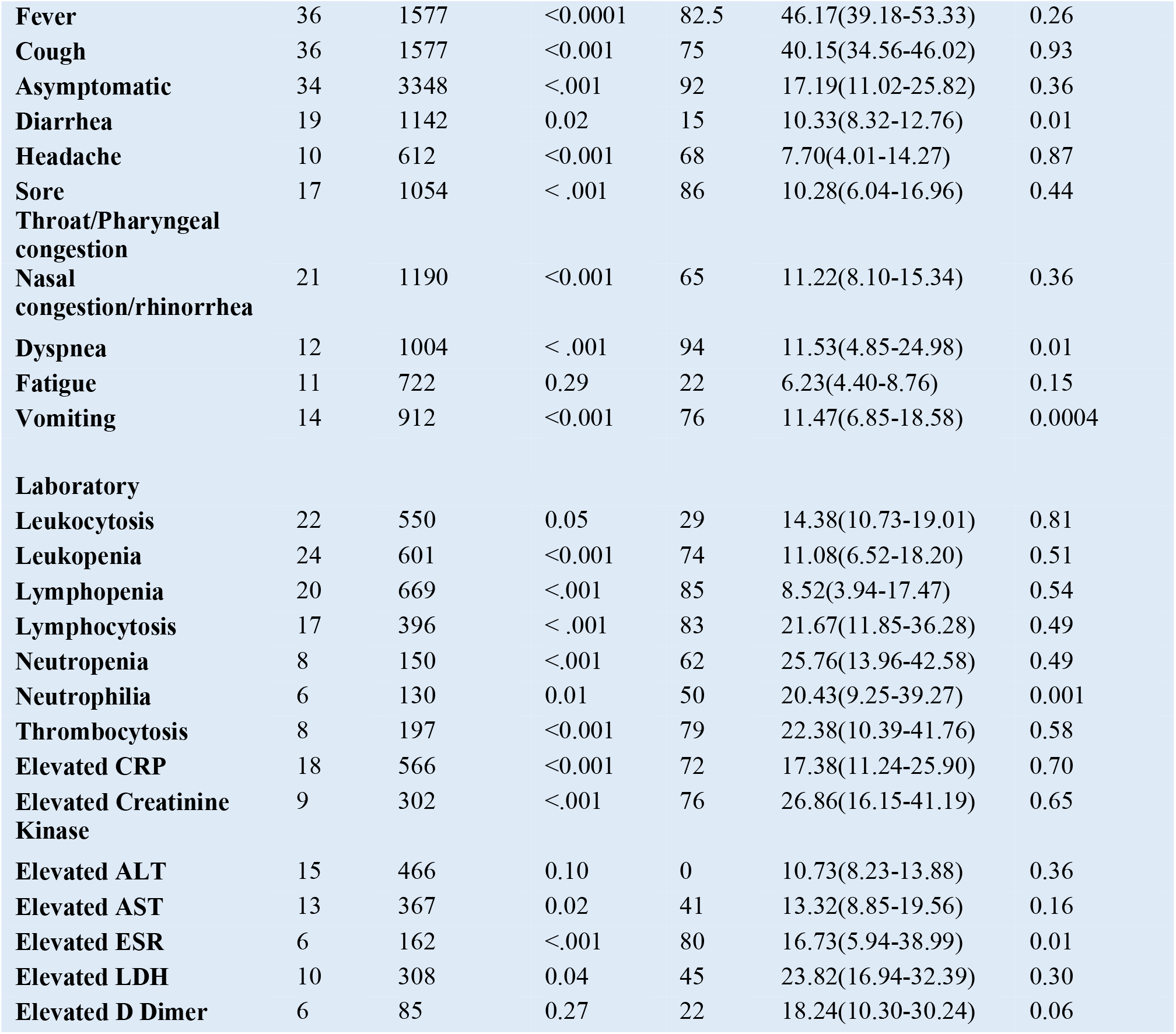
Pooled prevalence of clinical characteristic and laboratory findings in COVID-19 children using random effects meta-analysis

**Figure 1.**
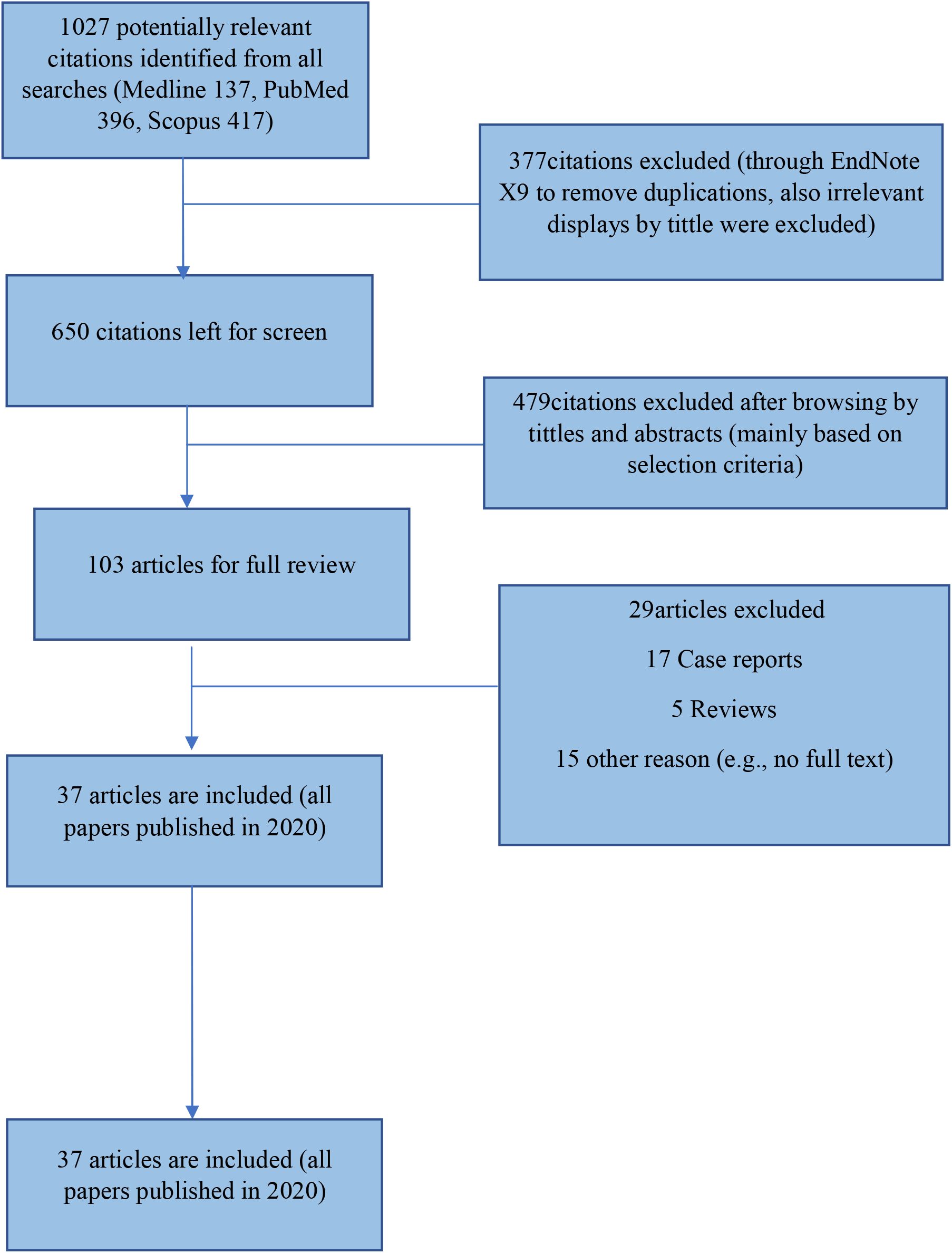
PRISMA flow diagram.

The estimated pooled prevalence of fever in COVID-19 children was 46.17% (95%CI 39.18-53.33%)(Figure2). The second most prevalent symptom prevalent in COVID-19 children was cough (40.15%, 95%CI 34.56-46.02%)(Figure3). An estimated 17.19% (95%CI 11.02-25.82%) of children did not have any prevalent symptoms and were asymptomatic. These symptoms were followed by less common symptoms of dyspnea (11.53%, 95% CI 4.85%-24.98%), vomiting (11.47%, 95%CI 6.85-18.58%), nasal congestion/rhinorrhea (11.22%,95%CI 8.10-15.34%), diarrhea (10.33%, 95%CI 8.32-12.76%), sore throat/pharyngeal congestion (10.28%, 95%CI 6.04-16.96%), headache (7.70%, 95%CI 4.01-14.27%) and fatigue (6.23%, 95%CI-4.40-8.76%) (Table 2).

**Figure 2.**
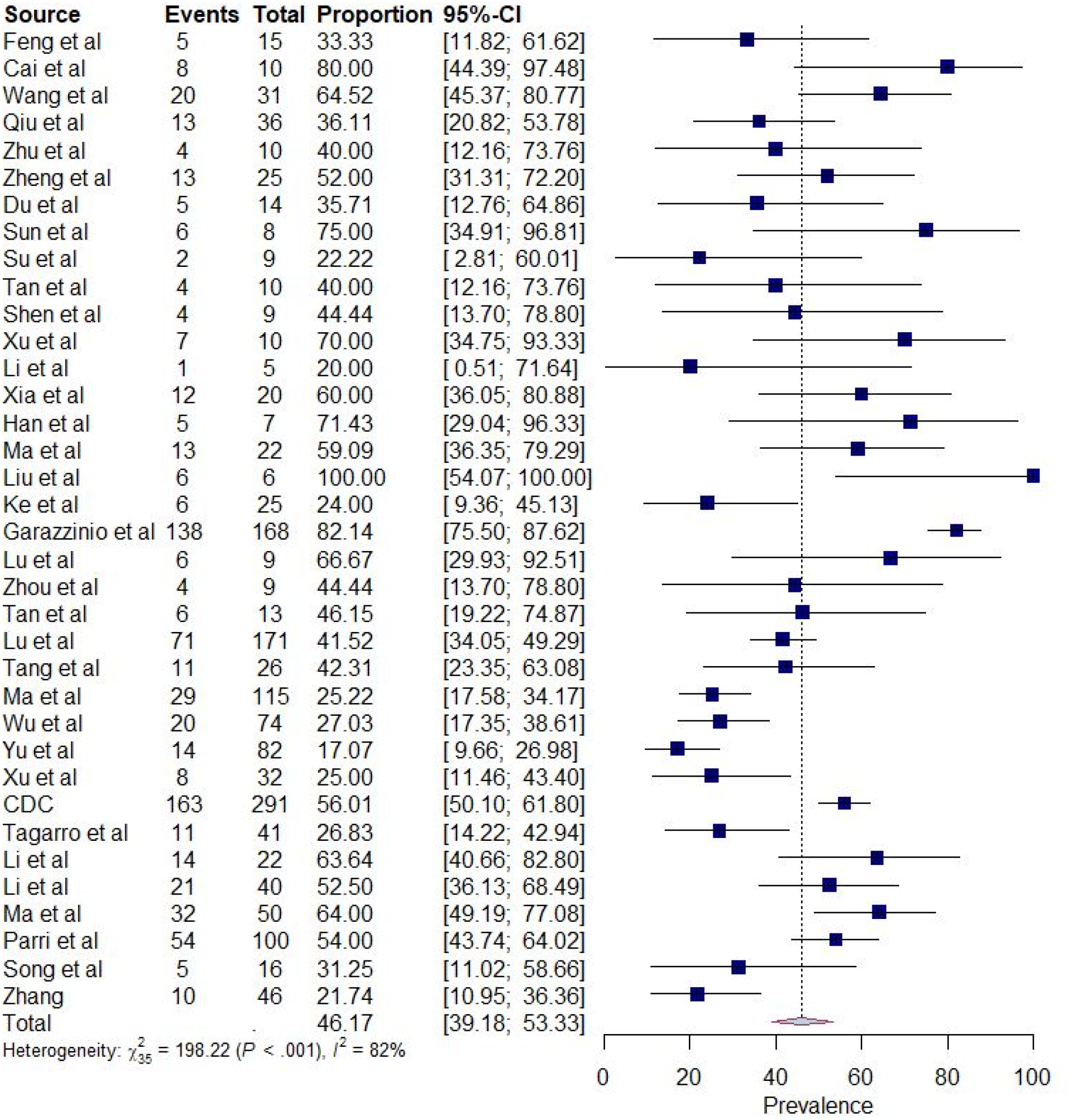
Prevalence of fever in COVID-19 positive children.

**Figure 3.**
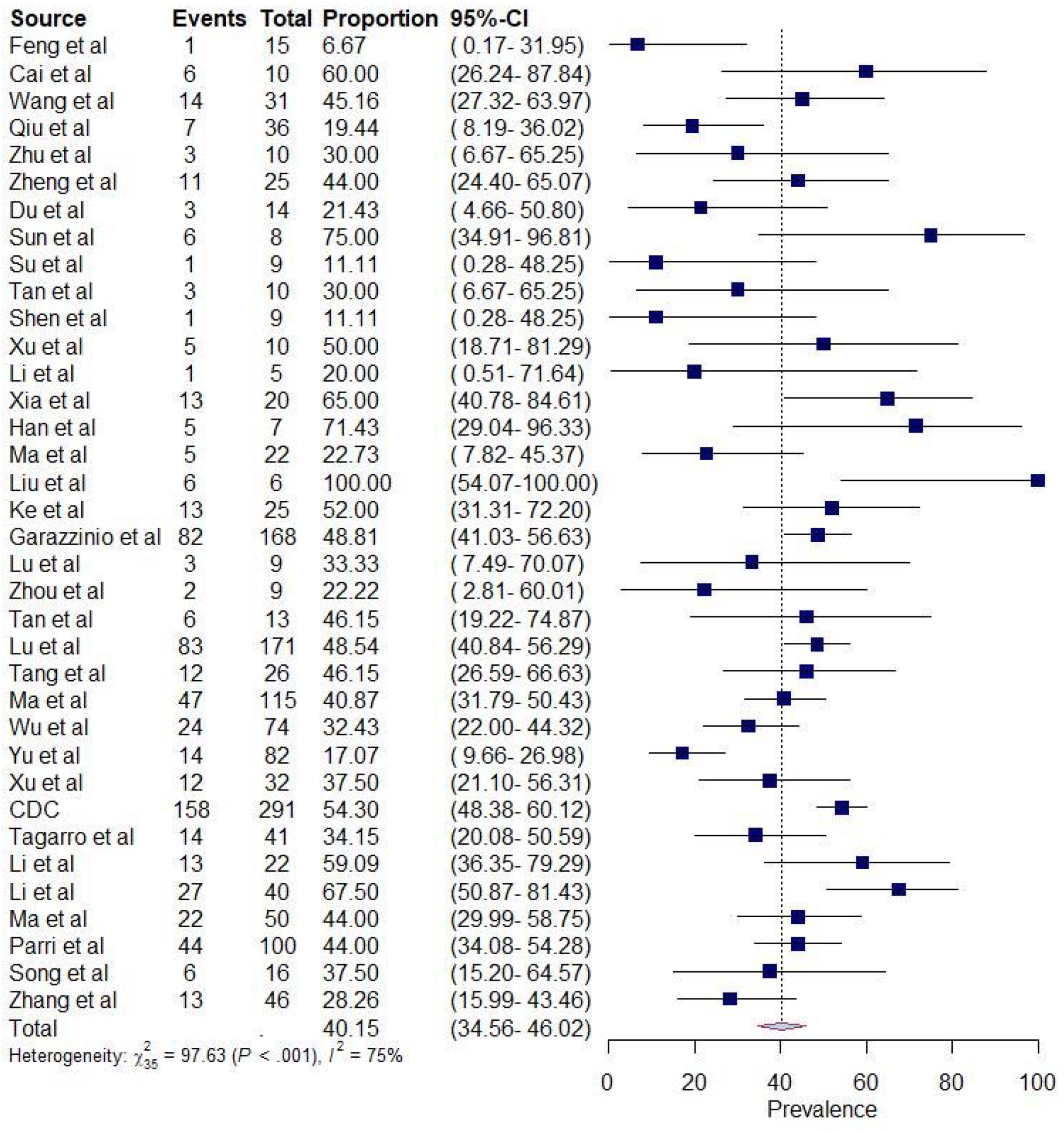
Prevalence of cough in COVID-19 positive children.

The most prevalent laboratory findings in COVID-19 children was elevated creatinine kinase (26.86%, 95%CI16.15-41.19%)(Figure4) and neutropenia (25.76%, 95%CI 13.96-42.58%)(Figure5). These were followed by elevated LDH (23.82%, 95%CI 16.94-32.39), thrombocytosis (22.38%, 95%CI 10.39-41.76%), lymphocytosis (21.67%, 95%CI 11.85-36.28%), neutrophilia (20.43%95%CI-9.25-39.27%), elevated D Dimer (18.24%, 95%CI 10.30-30.24%), elevated CRP (17.38%, 95%CI 11.24-25.90%), elevated ESR (16.73%,95%CI 5.94-38.99%), leukocytosis (14.38%.95%CI 10.73-19.01%), elevated AST (13.32%, 95%CI 8.85-19.56%), leukopenia (11.08%, 95%CI 6.52-18.20%), elevated ALT (10.73%, 95%CI 8.23-13.88%) and lymphopenia (8.52%, 95%CI 3.94-17.47%) in children with COVID-19. (Table2)

**Figure 4.**
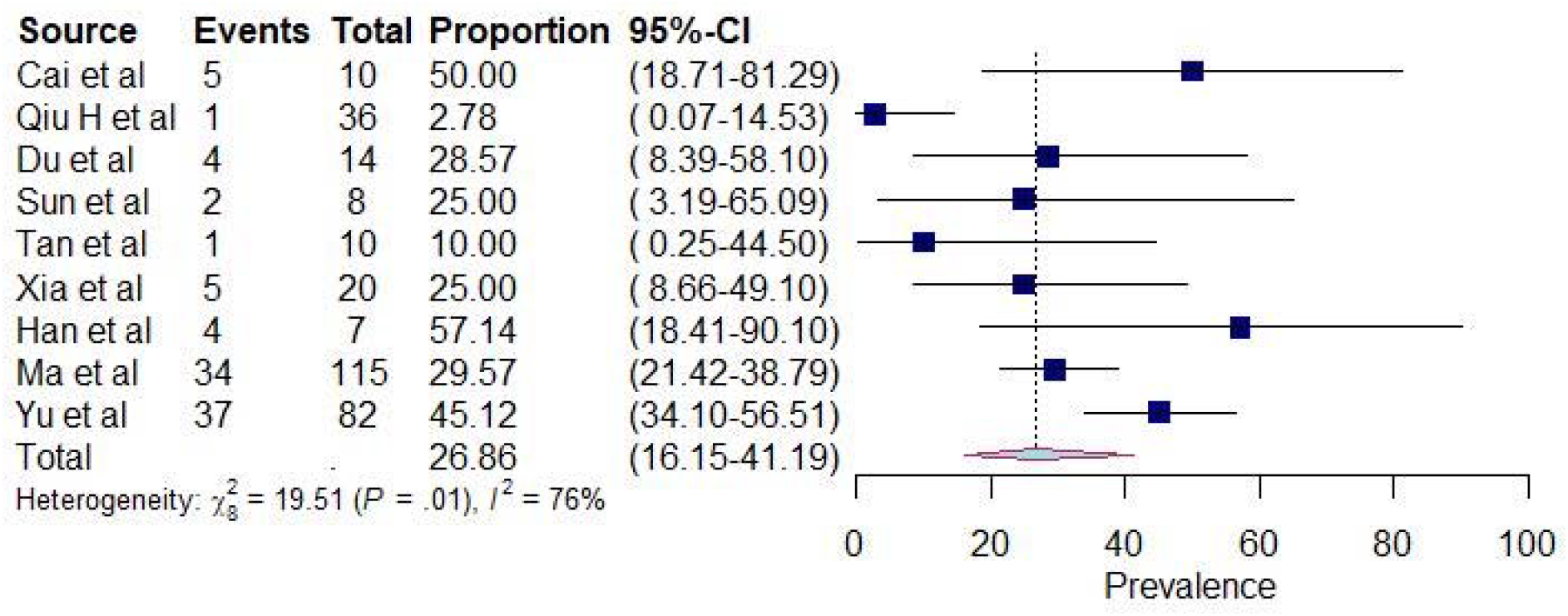
Prevalence of elevated creatinine kinase in COVID-19 positive children.

**Figure 5.**
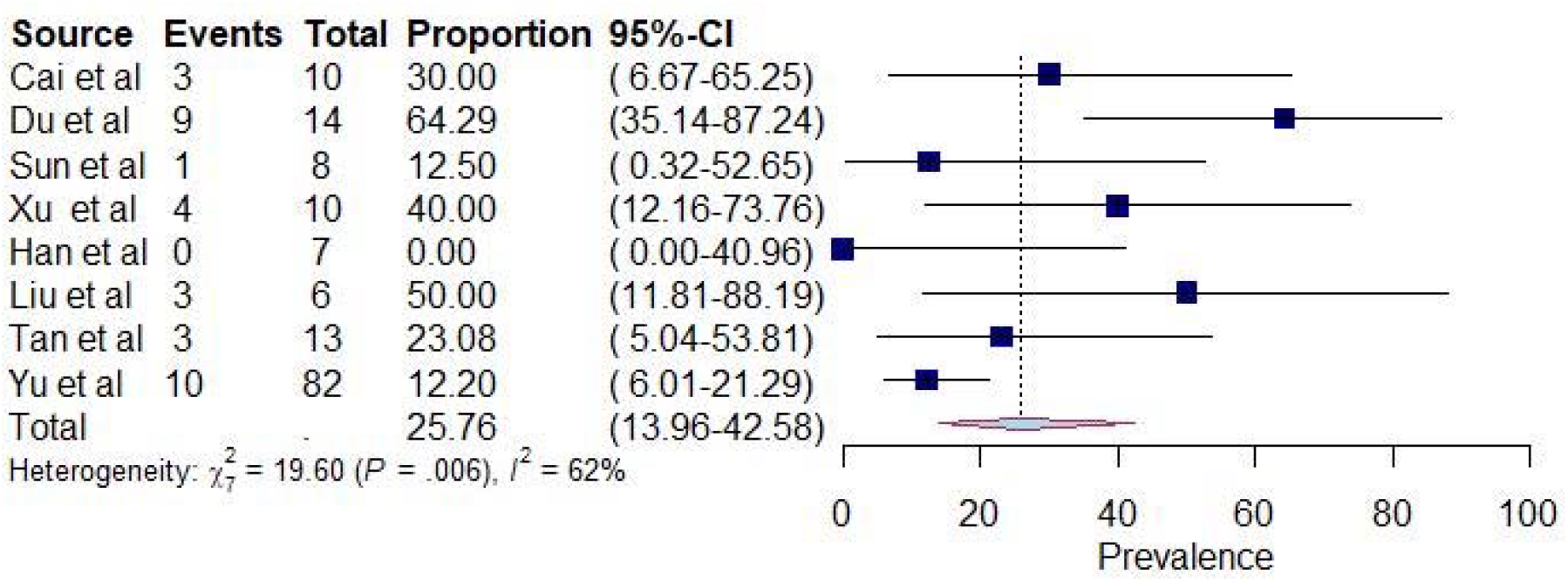
Prevalence of elevated neutropenia in COVID-19 positive children.

The *I*^*2*^ of the studies varied between 0 to 94%. Symptoms and laboratory findings with I^*2*^ greater than 50% and p < 0.05 were interpreted as having significant heterogeneity across studies. There was not any significant heterogeneity observed between studies for clinical symptoms and laboratory findings except for fatigue (p=0.29), elevated ALT (p=0.10) and elevated D Dimer (p=0.27) which was not explained by sample size and sex. (eTable 2 in the supplement)

There was no evidence of publication bias in the reporting of fever as a symptom, based on the observed funnel plot and Egger’s Test for funnel plot asymmetry (efigure20. in the Supplement) (Table 2) Similarly, there was no evidence of funnel plot asymmetry for other symptoms and laboratory findings except for clinical characteristic of diarrhea (p=0.01), vomiting (p=0.0004) and dyspnea (p=0.01) and laboratory findings of neutrophilia (p=0.001) and Elevated ESR9 (p=0.01) (Table 2).

## DISCUSSION

The pandemic caused by the outbreak of COVID-19 has led to a widespread catastrophic effect on global health. Adults are particularly susceptible to severe symptoms of COVID-19 as compared to children.(4-5) However, recent studies have shown increased incidence of Multisystemic Inflammatory Syndrome in Children (MIS-C) with COVID-19 infection. (14) According to the CDC, as of August 3, 7.3% of all the cases of COVID-19 were among children.(6) A cross-sectional study showed that 38% of the hospitalized children required intubation. (15) Therefore, in the current scenario the importance of understanding the presentation of COVID-19 in children should not be ignored.

This systematic review and meta-analysis summarizes the effect of COVID-19 on the pediatric population based on the available 24 studies published from January 1, 2020 to July 15, 2020 and included a total of 3712 male and female children. The predominant and most common symptom was fever which was present in 46.17% of the children diagnosed with COVID-19. 40.15% of children presented with cough, and 11.53% had difficulty breathing including tachypnea/dyspnea. 17.19% of the children were asymptomatic. Few children also presented with less common presentations including dyspnea, vomiting, nasal congestion/rhinorrhea, diarrhea, sore throat/pharyngeal congestion, headache, and fatigue. (Table2)

The most common hematological abnormality among children diagnosed with COVID-19 was elevated creatinine kinase and neutropenia which were both present in more than 25% of the children. Other common laboratory findings included elevated LDH, thrombocytosis, lymphocytosis, neutrophilia, elevated D Dimer, Elevated CRP, elevated ESR, leukocytosis, elevated AST and leukopenia (Table 2).

Diagnosing COVID-19 disease in children is a challenge since most of the children present asymptomatically or with flu-like symptoms which could occur with other viruses as well. Why children are less susceptible to COVID-19 also needs to be addressed.

Many different hypotheses have been proposed regarding the lower susceptibility of children compared to adults of COVID-19. According to one hypothesis, children are not able to mount as heightened of an immune response compared to adults, thus incurring less organ damage. This might be due to immature immune system in children which is drastically changing after birth and is different as compared to adults in their function, immunological response, and their composition. (7, 16) The preliminary analysis conducted by Wong showed that children with COVID-19 had lower cytokine levels.(17)

Another hypothesis states that high viral loads of SARS-CoV-2 virus are associated with more severe symptoms of the disease. (18) Therefore, presence of other viruses in the mucosa of lungs and airways of young children could limit the growth of SARS-CoV-2 by direct virus to virus interactions and competition leading to milder disease as compared to adults. (7, 18) Children often get upper respiratory infection from other coronaviruses in the community, which might be providing protection against SARS-CoV-2.(19)

It has been observed that COVID-18 is associated with severe illness in adults who have chronic underlying comorbidities like hypertension, diabetes mellitus and use ACE inhibitor drugs/ARB drugs.(20) It is hypothesized that these drugs lead to increased expression of ACE 2 receptors; since SARS-CoV-2 attacks the ACE-2 receptors, this could lead to increased severity. (20) However, since children generally do not require these drugs, ACE2 receptor expression may differ in children, possibly leading them to experience a milder form of COVID-19 disease.(7, 20-22) Smoking is another factor which leads to an increases the risk of severity in adults.(7, 21, 22)

Underlying comorbidities in children may serve as a risk factor for severe COVID-19 illness and hospitalization requiring respiratory support i.e., tracheostomy.(15) The most common underlying comorbidities are congenital conditions, chronic pulmonary diseases, immunosuppression due to cancer, chemotherapy, obesity, hematopoietic cell or solid organ transplant and high doses of glucocorticoids.(15)

A strength of our study is that a comprehensive search strategy was followed with strict inclusion criteria. However, our study also has several limitations. The majority of the included studies were conducted in China and some studies did not report laboratory findings. Some of the studies also had small sample sizes. We found evidence of publication bias for several of the outcomes. These outcomes tended to have a lower estimated pooled prevalence, and it is possible that some studies did not collect or report data on symptoms they perceived as rare. COVID-19 is characterized by a wide variety of symptoms and our understanding of this has evolved considerably over time. It has taken longer to detect less common symptoms such as gastrointestinal issues like vomiting and diarrhea. Studies with a higher proportion of asymptomatic individuals may have also collected data on fewer symptoms. However, we did not find evidence of publication bias in the majority of symptoms and laboratory findings included in this study.

More research studies need to be conducted to determine the possible mechanism of the mild form of COVID-19 disease in children. Further studies also need to be conducted to determine the rate of transmission by COVID-19 children especially asymptomatic children.

## CONCLUSION

Children with COVID-19 present with a varied range of symptoms. The predominant symptoms in children are fever and cough followed by dyspnea, vomiting, nasal congestion/rhinorrhea, diarrhea, sore throat/pharyngeal congestion, headache, and fatigue. Many children also present asymptomatically. The most common hematological abnormality among children diagnosed with COVID-19 was elevated creatinine kinase and neutropenia which was present in more than 25% of the children. Other common laboratory findings included elevated LDH, thrombocytosis, lymphocytosis, neutrophilia, elevated D Dimer, Elevated CRP, elevated ESR, leukocytosis, elevated AST, and leukopenia. There was a low prevalence of elevated ALT and lymphopenia.

Recently, we have seen some COVID-19 positive children experiencing critical illness such as Multisystem inflammatory syndrome in children (MIS-C). As COVID-19 is a new disease and we still lack information about its long-term effects, further studies are needed to understand the complete picture of the COVID-19 in children.

## Data Availability

Studies referred to in the manuscript are available freely in pubmed.

## Author Contributions

Dr.Harmeet Kharoud had full access to all of the data in the study and takes responsibility for the integrity of the data and the accuracy of data analysis.

Concept and Design: Dr.Harmeet Kaur Kharoud and Dr.Gagan Deep Singh

Acquisition, analysis or interpretation of data: Dr.Harmeet Kaur Kharoud, Dr.Rizwana Asim and Dr.Gagan Deep Singh, Lovepreet Chahal

Drafting of the manuscript: Dr. Harmeet Kaur Kharoud, Dr. Rizwana Asim, Lovepreet Chahal

Critical revision of the manuscript for important intellectual content: Dr. Gagan Deep Singh, Dr.Harmeet Kaur Kharoud, Dr.Rizwana Asim, Lianne Siegel.

Statistical Analysis: Dr. Harmeet Kharoud, Lianne Siegel

Supervision: Dr. Harmeet Kharoud, Dr. Gagan Deep Singh.

Additional Contributions-Raj Mehta assisted in statistical analysis and assisted in revision of manuscript. Mandakinee Singh Patel assisted in statistical analysis. All are affiliated with the University of Minnesota. Sarbrinder Dhillon helped in critical review of the manuscript. He is affiliated with Saba University.

Funding sources

NIH NHLBI T32HL129956 (LS)

Authors declare no conflict of interest.

## Notes

### Competing Interest Statement

The authors have declared no competing interest.

### Author Declarations

Since this is a meta-analysis IRB approval was not required

## References

1. (CDC) CfDCaP. Coronavirus Disease 2019 (COVID-19).

2. Du W, Yu J, Wang H, Zhang X, Zhang S, Li Q, et al. Clinical characteristics of COVID-19 in children compared with adults in Shandong Province, China. Infection. 2020;48(3):445–52.

3. Su L, Ma X, Yu H, Zhang Z, Bian P, Han Y, et al. The different clinical characteristics of corona virus disease cases between children and their families in China - the character of children with COVID-19. Emerg Microbes Infect. 2020;9(1):707–13.

4. Ludvigsson JF. Systematic review of COVID-19 in children shows milder cases and a better prognosis than adults. Acta Paediatr. 2020;109(6):1088–95.

5. Dong Y, Mo X, Hu Y, Qi X, Jiang F, Jiang Z, et al. Epidemiology of COVID-19 Among Children in China. Pediatrics. 2020;145(6).

6. (CDC) CfDCaP. Care For Children.

7. Brodin P. Why is COVID-19 so mild in children? Acta Paediatrica. 2020;109(6):1082–3.

8. Moher D, Liberati A, Tetzlaff J, Altman DG, Group P. Preferred reporting items for systematic reviews and meta-analyses: the PRISMA statement. PLoS Med. 2009;6(7):e1000097.

9. von Elm E, Altman DG, Egger M, Pocock SJ, Gotzsche PC, Vandenbroucke JP, et al. The Strengthening the Reporting of Observational Studies in Epidemiology (STROBE) statement: guidelines for reporting observational studies. Lancet. 2007;370(9596):1453–7.

10. Viechtbauer W. Conducting Meta-Analyses in R with the metafor Package. 2010. 2010;36(3):48.

11. Lin L, Chu H. Meta-analysis of Proportions Using Generalized Linear Mixed Models. Epidemiology. 2020;31(5):713–7.

12. Higgins JP, Thompson SG, Deeks JJ, Altman DG. Measuring inconsistency in meta-analyses. BMJ. 2003;327(7414):557–60.

13. Higgins JPT, Thompson SG. Quantifying heterogeneity in a meta-analysis. Statistics in Medicine. 2002;21(11):1539–58.

14. Cheung EW, Zachariah P, Gorelik M, Boneparth A, Kernie SG, Orange JS, et al. Multisystem Inflammatory Syndrome Related to COVID-19 in Previously Healthy Children and Adolescents in New York City. JAMA. 2020.

15. Shekerdemian LS, Mahmood NR, Wolfe KK, Riggs BJ, Ross CE, McKiernan CA, et al. Characteristics and Outcomes of Children With Coronavirus Disease 2019 (COVID-19) Infection Admitted to US and Canadian Pediatric Intensive Care Units. JAMA Pediatrics. 2020.

16. Simon AK, Hollander GA, McMichael A. Evolution of the immune system in humans from infancy to old age. Proc Biol Sci. 2015;282(1821):20143085.

17. Mallapaty S. How do children spread the coronavirus? The science still isn’t clear. Nature. 2020;581(7807):127–8.

18. Liu Y, Yan LM, Wan L, Xiang TX, L.A, Liu JM, et al. Viral dynamics in mild and severe cases of COVID-19. Lancet Infect Dis. 2020;20(6):656–7.

19. Dhochak N, Singhal T, Kabra SK, Lodha R. Pathophysiology of COVID-19: Why Children Fare Better than Adults? Indian J Pediatr. 2020;87(7):537–46.

20. Diaz JH. Hypothesis: angiotensin-converting enzyme inhibitors and angiotensin receptor blockers may increase the risk of severe COVID-19. J Travel Med. 2020;27(3).

21. Ferrario CM, Jessup J, Chappell MC, Averill DB, Brosnihan KB, Tallant EA, et al. Effect of Angiotensin-Converting Enzyme Inhibition and Angiotensin II Receptor Blockers on Cardiac Angiotensin-Converting Enzyme 2. Circulation. 2005;111(20):2605–10.

22. van Zyl-Smit RN, Richards G, Leone FT. Tobacco smoking and COVID-19 infection. The Lancet Respiratory Medicine.

23. Feng K, Yun YX, Wang XF, Yang GD, Zheng YJ, Lin CM, et al. [Analysis of CT features of 15 Children with 2019 novel coronavirus infection]. Zhonghua Erke Zazhi.58(0):E007.

24. Cai J, Xu J, Lin D, Yang Z, Xu L, Qu Z, et al. A Case Series of children with 2019 novel coronavirus infection: clinical and epidemiological features. Clin Infect Dis. 2020.

25. Wang D, Ju XL, Xie F, Lu Y, Li FY, Huang HH, et al. [Clinical analysis of 31 cases of 2019 novel coronavirus infection in children from six provinces (autonomous region) of northern China]. Zhonghua Erke Zazhi.58(4):E011.

26. Qiu H, Wu J, Hong L, Luo Y, Song Q, Chen D. Clinical and epidemiological features of 36 children with coronavirus disease 2019 (COVID-19) in Zhejiang, China: an observational cohort study. The Lancet Infectious Diseases. 2020;25:25.

27. Zhu L, Wang J, Huang R, Liu L, Zhao H, Wu C, et al. Clinical characteristics of a case series of children with coronavirus disease 2019. Pediatric Pulmonology. 2020;08:08.

28. Zheng F, Liao C, Fan QH, Chen HB, Zhao XG, Xie ZG, et al. Clinical Characteristics of Children with Coronavirus Disease 2019 in Hubei, China. Current Medical Science. 2020;24:24.

29. Du W, Yu J, Wang H, Zhang X, Zhang S, Li Q, et al. Clinical characteristics of COVID-19 in children compared with adults in Shandong Province, China. Infection. 2020;16:16.

30. Sun D, Li H, Lu XX, Xiao H, Ren J, Zhang FR, et al. Clinical features of severe pediatric patients with coronavirus disease 2019 in Wuhan: a single center’s observational study. World Journal of Pediatrics. 2020;19:19.

31. Su L, Ma X, Yu H, Zhang Z, Bian P, Han Y, et al. The different clinical characteristics of corona virus disease cases between children and their families in China – the character of children with COVID-19. Emerging Microbes & Infections.9(1):707–13.

32. Tan YP, Tan BY, Pan J, Wu J, Zeng SZ, Wei HY. Epidemiologic and clinical characteristics of 10 children with coronavirus disease 2019 in Changsha, China. Journal of Clinical Virology. 2020;127.

33. Shen Q, Guo W, Guo T, Li J, He W, Ni S, et al. Novel coronavirus infection in children outside of Wuhan, China. Pediatric Pulmonology. 2020;07:07.

34. Xu Y, Li X, Zhu B, Liang H, Fang C, Gong Y, et al. Characteristics of pediatric SARS-CoV-2 infection and potential evidence for persistent fecal viral shedding. Nature Medicine.26(4):502–5.

35. Li W, Cui H, Li K, Fang Y, Li S. Chest computed tomography in children with COVID-19 respiratory infection. Pediatric Radiology. 2020;11:11.

36. Xia W, Shao J, Guo Y, Peng X, Li Z, Hu D. Clinical and CT features in pediatric patients with COVID-19 infection: Different points from adults. Pediatr Pulmonol. 2020;55(5):1169–74.

37. Han YN, Feng ZW, Sun LN, Ren XX, Wang H, Xue YM, et al. A comparative-descriptive analysis of clinical characteristics in 2019-coronavirus-infected children and adults. Journal of Medical Virology. 2020;06:06.

38. Ma H SJ, Wang Y, Zhai A, Zheng N, Li Q et al. High resolution CT features of COVID-19 in children. Chinese Journal of Radiology (China) 2020 Apr 10;54(4).

39. Bai K, Liu W, Liu C, Fu Y, Hu J, Qin Y, et al. Clinical Analysis of 25 Novel Coronavirus Infections in Children. Pediatr Infect Dis J. 2020.

40. Garazzino S, Montagnani C, Donà D, Meini A, Felici E, Vergine G, et al. Multicentre Italian study of SARS-CoV-2 infection in children and adolescents, preliminary data as at 10 April 2020. Eurosurveillance. 2020;25(18):2000600.

41. Lu Y, Wen H, Rong D, Zhou Z, Liu H. Clinical characteristics and radiological features of children infected with the 2019 novel coronavirus. Clinical Radiology. 2020.

42. Zhou Y, Yang GD, Feng K, Huang H, Yun YX, Mou XY, et al. [Clinical features and chest CT findings of coronavirus disease 2019 in infants and young children]. Zhongguo Dang Dai Er Ke Za Zhi. 2020;22(3):215–20.

43. Tan X, Huang J, Zhao F, Zhou Y, Li JQ, Wang XY. [Clinical features of children with SARS-CoV-2 infection: an analysis of 13 cases from Changsha, China]. Zhongguo Dang Dai Er Ke Za Zhi. 2020;22(4):294–8.

44. Lu X, Zhang L, Du H, Zhang J, Li YY, Qu J, et al. SARS-CoV-2 Infection in Children. N Engl J Med. 2020;382(17):1663–5.

45. Tang A, Xu W, shen m, Chen P, Li G, Liu Y, et al. A retrospective study of the clinical characteristics of COVID-19 infection in 26 children. medRxiv. 2020:2020.03.08.20029710.

46. Ma H, Hu J, Tian J, Zhou X, Li H, Laws MT, et al. A single-center, retrospective study of COVID-19 features in children: a descriptive investigation. BMC Med. 2020;18(1):123.

47. Wu Q, Xing Y, Shi L, Li W, Gao Y, Pan S, et al. Epidemiological and Clinical Characteristics of Children with Coronavirus Disease 2019. medRxiv. 2020:2020.03.19.20027078.

48. Yu H, Cai Q, Dai X, Liu X, Sun H. The clinical and epidemiological features and hints of 82 confirmed COVID-19 pediatric cases aged 0-16 in Wuhan, China. medRxiv. 2020:2020.03.15.20036319.

49. Xu H, Liu E, Xie J, Smyth R, Zhou Q, Zhao R, et al. A follow-up study of children infected with SARS-CoV-2 from Western China. medRxiv. 2020:2020.04.20.20073288.

50. Team CC-R. Coronavirus Disease 2019 in Children – United States, February 12-April 2, 2020. MMWR Morb Mortal Wkly Rep. 2020;69(14):422–6.

51. Tagarro A, Epalza C, Santos M, Sanz-Santaeufemia FJ, Otheo E, Moraleda C, et al. Screening and Severity of Coronavirus Disease 2019 (COVID-19) in Children in Madrid, Spain. JAMA Pediatr. 2020.

52. Li B, Shen J, Li L, Yu C. Radiographic and Clinical Features of Children With Coronavirus Disease (COVID-19) Pneumonia. Indian Pediatr. 2020;57(5):423–6.

53. Li H, Chen K, Liu M, Xu H, Xu Q. The profile of peripheral blood lymphocyte subsets and serum cytokines in children with 2019 novel coronavirus pneumonia. J Infect. 2020;81(1):115–20.

54. MA Yao-Ling XS-Y, WANG Min, ZHANG Si-Min, DU Wen-Hui, CHEN Qiong. Clinical features of children with SARS-CoV-2 infection: an analysis of 115 cases. CJCP. 2020;22(4):290–3.

55. Parri N, Lenge M, Buonsenso D. Children with Covid-19 in Pediatric Emergency Departments in Italy. New England Journal of Medicine. 2020;383(2):187–90.

56. Song W, Li J, Zou N, Guan W, Pan J, Xu W. Clinical features of pediatric patients with coronavirus disease (COVID-19). J Clin Virol. 2020;127:104377.

57. Zhang B, Liu S, Zhang J, Xiao J, Zhu S, Dong Y, et al. Children hospitalized for coronavirus disease 2019 (COVID-19): A multicenter retrospective descriptive study. J Infect. 2020;81(2):e74–e5.

